# Influence of immunosuppressive regimen on diffusivity and oxygenation of kidney transplants – analysis of functional MRI data from the randomized ZEUS trial

**DOI:** 10.1101/2022.04.11.22273639

**Authors:** Laila-Yasmin Mani, Jasna Cotting, Bruno Vogt, Ute Eisenberger, Peter Vermathen

## Abstract

The ZEUS study was a multi-center randomized controlled trial investigating the effect of an early conversion from ciclosporin-based to an everolimus-based regimen on graft function twelve months post-transplantation. In this investigator-initiated sub-study, functional magnetic resonance imaging (fMRI) of kidney grafts was prospectively performed to non-invasively assess differences in graft oxygenation, diffusion and perfusion between groups and time-points using diffusion-weighted imaging (DWI) and blood oxygen level-dependent (BOLD)-MRI. Sixteen patients underwent DWI and BOLD-MRI at months 4.5 and 12 post-transplantation on a 3 Tesla and 1.5 Tesla (n=3) MR scanner. After exclusion due to image quality, outlier values or missing data, DWI was analyzed in ten, BOLD in eight subjects. The diffusion coefficient ADC_D_ decreased in the CsA-group over time, whereas it increased in the EVE-group (*p*=0.046, medulla). The change in ADC_D_ from month 4.5 to 12 significantly differed between groups in cortex (*p*=0.033) and medulla (*p*=0.019). In BOLD, cortico-medullary transverse relaxation rate R2* increased (decreased tissue oxygen) in the CsA-treated and decreased in the EVE-treated group over time. Similarly, R2* values at month 12 were higher in the CsA- vs. EVE-treated group. There was no significant difference for the perfusion fraction F_P_. In conclusion, this prospective sub-study of the ZEUS trial suggests an impact of immunosuppressive regimen on fMRI parameters of the kidney graft.

## 1. Introduction

Kidney transplantation is the treatment of choice for patients with end-stage kidney disease prolonging survival [1]. Thanks to highly efficient immunosuppressive therapies, a dramatic reduction in the risk of allograft rejection has been achieved since its early period. Until now, the mainstay of post-transplant immunosuppressive regimens are calcineurin inhibitors acting on T cell and T cell-dependent B cell activation. However, major adverse effects including nephrotoxicity, arterial hypertension and de novo diabetes mellitus limit their usefulness in kidney transplant recipients [2]. Hence, existing efforts target therapeutic strategies maximally limiting the exposition to calcineurin inhibitors either by dose reduction, by shortening of administration period or by replacement by other drugs [3-5].

The ZEUS study was a multi-center open-label randomized controlled trial designed to investigate the effect of an early conversion from a calcineurin inhibitor-based immunosuppression with ciclosporin (CsA) to a mammalian-target-of-rapamycin inhibitor-based regimen with everolimus (EVE) on graft function twelve months post-transplantation [6]. An improvement of the estimated glomerular filtration rate (eGFR) at twelve months was demonstrated in the EVE-treated as compared to the CsA-treated group, which persisted five years post-transplant, despite a non-significantly higher number of rejection episodes [6, 7].

Functional magnetic resonance imaging (fMRI) techniques represent an attractive diagnostic tool for renal investigations allowing the non-invasive simultaneous assessment of several aspects of kidney function such as tissue structure, perfusion and oxygenation in addition to morphological imaging, without need for contrast media [8, 9]. Diffusion-weighted imaging (DWI) evaluates organ diffusivity and microperfusion, while blood oxygen level dependent (BOLD)-MRI assesses tissue oxygenation. Whereas DWI has been investigated as a non-invasive marker of kidney function, BOLD has proven particularly useful to study acute and chronic effects of various interventions [9-12].

The purpose of this investigator-initiated local sub-study of the ZEUS trial was the evaluation of additional aspects of kidney graft function according to the immunosuppressive regimen using fMRI methods. With the exception of one study performing BOLD after acute intake of ciclosporin, this question had not been addressed so far to the best of our knowledge [13]. Due to the known acute and chronic effects of calcineurin inhibitor nephrotoxicity including vasoconstriction of afferent arterioles and development of interstitial fibrosis and tubular atrophy (IFTA), we hypothesized that diffusivity and microperfusion as measured by DWI as well as tissue oxygenation measured by BOLD-MRI of kidney grafts differ between patients treated with ciclosporin as opposed to everolimus [14]. The results show an improvement of graft diffusivity and a tendency for ameliorated tissue oxygenation after switch to everolimus as compared to patients maintained on ciclosporin.

## 2. Materials and Methods

The protocol of the present investigator-initiated prospective single-center sub-study of the ZEUS trial was approved by the local ethics committee (Canton of Bern, Switzerland, approval number 2004/213) and conducted in accordance with the Declarations of Helsinki and Istanbul [15, 16].

### 2.1. Study population

All kidney transplant recipients included in the ZEUS trial from our study center were eligible for the current study [6]. Specific exclusion criteria were lack of consent to participate in the sub-study, body weight >200 kg, classical contraindications to MRI as well as implanted metallic material without prior 3T-MRI after implantation. Among the 300 study participants of the ZEUS study, 37 patients had been enrolled from our center and represented the screening population for this sub-study.

### 2.2. Study design

The ZEUS study was a 12-month multi-center randomized controlled parallel-group trial, the protocol of which has been published previously [6]. Patients were screened among participants of the ZEUS trial before randomization at the outpatient University Clinic for Nephrology and Hypertension in Bern (Figure 1). Written informed consent was obtained from each participant prior to inclusion. The two study visits took place at baseline (4.5 months after transplantation, before randomization within the ZEUS trial) as well as at month 12 after transplantation. A light meal was allowed on the study day. Laboratory tests including serum creatinine, serum urea, CsA trough levels and urinary protein as well as ambulatory blood pressure measurement (ABPM), duplex ultrasound scan and functional MRI of the kidney graft were performed at both visits. Baseline clinical characteristics based on medical chart review and EVE trough level were determined at baseline- and 12 month-visits respectively. In addition, protocol transplant biopsies were performed at baseline and 12 months as part of this study amendment.

**Figure 1.**
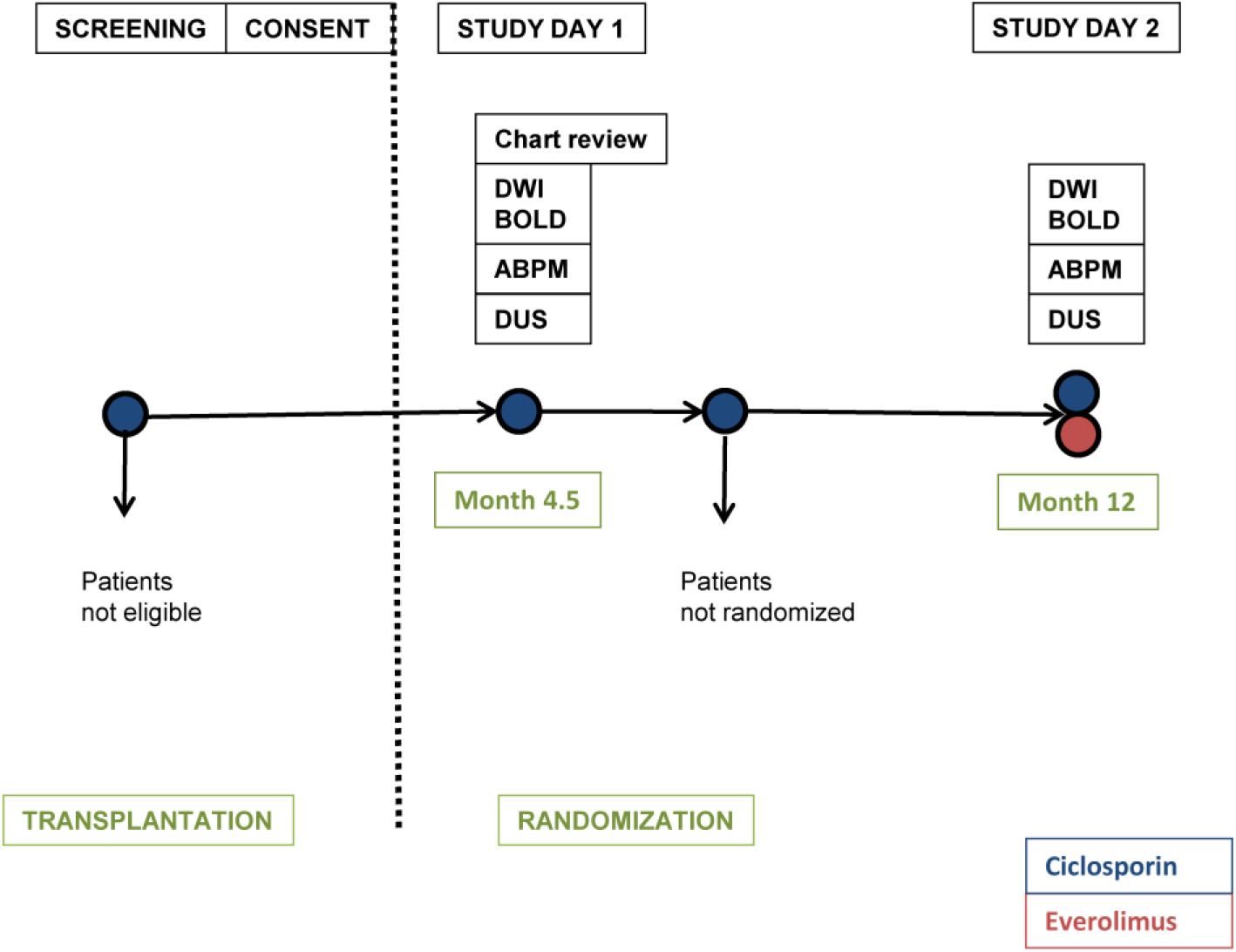
Study design. DWI (diffusion-weighted imaging); BOLD (blood oxygen level dependent-MRI); ABPM (ambulatory blood pressure measurement); DUS (duplex ultrasound scan).

### 2.3. MRI protocol

MRI data were acquired on a 3.0 T whole body MR Scanner (Tim Trio®; Siemens Healthcare, Erlangen, Germany) for all subjects but three who underwent MRI on a 1.5 T MR scanner (Sonata®; Siemens Healthcare, Erlangen, Germany). During each of the two MRI sessions anatomical MRI, DWI and BOLD-MRI were performed.

Intravoxel incoherent motion (IVIM)-DWI yielded the perfusion-cleared apparent diffusion coefficient ADC_D_ and the perfusion fraction F_P_. Coronal multisection echoplanar DWI was performed with the following parameters: 11 slices (thickness: 5mm, intersection gap: 1mm), field of view (FOV)=400×400mm2, matrix=128×128, six averages, bandwidth=2300Hz/pixel and partial Fourier 6/8. Ten diffusion gradient b-values were applied (in sec/mm2): b=0, 10, 20, 50, 100, 180, 300, 420, 550, and 700. The gradients were applied in three orthogonal directions and subsequently averaged, minimizing effects of diffusion anisotropy. Parallel imaging (iPAT, mSENSE) with a reduction factor of 2 was applied. A TE of 64msec was used. Acquisition time for DWI was 8:06min.

BOLD-MRI takes advantage of deoxygenated hemoglobin as an endogenous contrast agent, which influences the relaxation time T2^*^ yielding the transverse relaxation rate R2^*^ (equal to 1/T2^*^) which correlates to tissue oxygen content provided that confounding factors such as blood volume or hydration state are excluded [8, 9]. For BOLD-MRI, a multiple gradient recalled echo sequence (mGre) was used. Four to six coronal slices were acquired with a slice thickness of 5mm and an intersection gap of 1mm, a FOV of 400×400mm2, a matrix size of 256×256, on average. Other parameters were: TR of 65msec, TE of 6–52msec, inter-echo spacing time of 4.2msec, flip angle of 30°, and bandwidth was 330Hz/pixel. Twelve T2*-weighted images, corresponding to 12 different echoes, were acquired for each slice within a single breath-hold of 17 sec.

A maximum of three regions of interest (ROIs) traced in medulla and cortex were analyzed in every slice (BOLD: 4-6, DWI: 4 slices) for each medulla and cortex. ROIs were manually defined by the same blinded investigator on images handed over in a random fashion. ROIs were traced in the medulla and cortex. Data were analyzed using in-house custom-scripts written in IDL® and MATLAB®. The obtained values were read into MS Excel® for further statistical processing. GraphPad Prism 9® was used for figure preparation.

### 2.4. Duplex ultrasound scan

Duplex ultrasound scan was performed at both study visits by the same experienced nephrologist on a Siemens Acuson Sequoia 512 machine. Recorded parameters were resistive indices (RI) measured at <u>intralobar</u> arteries (as a mean of superior, median and lower).

### 2.5. Ambulatory blood pressure measurement

Ambulatory blood pressure measurement was performed at both study visits using a Profilomat II® device (Disetronic Medical Systems, Burgdorf, Switzerland). Recorded variables were the overall mean arterial systolic, diastolic and mean pressures and dipping effect (in mmHg).

### 2.6 Laboratory analyses

Laboratory analyses were performed according to the ZEUS study protocol as specified in section 2.2 in the central laboratory of the Bern University Hospital. Glomerular filtration rate was estimated (eGFR) according to chronic kidney disease epidemiology 2021 formula.

### 2.7. Histopathological analysis of kidney grafts

Histopathological analysis of renal tissue obtained from protocol graft biopsies was performed according to clinical routine in the department of pathology of the University of Bern. Biopsy reports were retrospectively assessed for the presence of IFTA, arteriolar hyalinosis and rejection in a semi-quantitative manner.

### 2.8. Outcome measures

Primary outcome were the difference between the CsA-treated vs EVE-treated patient groups in ADC_D_, F_P_, R2* and cortico-medullary ratios (MCR) MCR ADC_D_, MCR F_P_ and MCR R2* at month 12 as well as in the change of ADC_D,_ F_P_ and R2* and the MCR from month 4.5 to month 12. Secondary outcomes were the change in ADC_D_, F_P_, R2* and cortico-medullary ratios from month 4.5 to month 12 in each medication group; the difference in mean RI and overall mean systolic, diastolic and mean blood pressure and dipping values between time-points for medication groups and the difference in changes from baseline to month 12 between groups; the correlation of fMRI parameters and their changes over time across and within medication groups with eGFR, RI, overall mean systolic, diastolic and mean blood pressure as well as dipping values.

### 2.9. Statistical analysis

Continuous variables were expressed as means with standard deviation (SD) or medians with range between minimal and maximal value. Wilcoxon-signed-rank-test was used for cortico-medullary differences and longitudinal changes in fMRI, RI and ABPM parameters within groups. Mann-Whitney-U-test was performed for comparisons across groups of absolute fMRI, RI and ABPM values as well as of longitudinal changes in these parameters. Correlations between fMRI absolute values, changes over time and differences across groups with eGFR, medication levels, RI and ABPM were determined by Kendall’s Tau correlation coefficient analysis. Data were analyzed using IBM SPSS Statistics 24® and MS Office 2007®.

## 3. Results

### 3.1. Study population

#### 3.1.1. Patient enrolment

From September 2005 until December 2006, 70 patients received a kidney transplant in our center and were screened at the outpatient University Clinic for Nephrology and Hypertension in Bern for participation in the ZEUS trial and the MRI substudy. A total of 37 subjects were enrolled in the ZEUS trial, 16 of which were included in the current study and underwent MR measurements. One patient was subsequently excluded because of lack of randomization (Figure 2).

**Figure 2.**
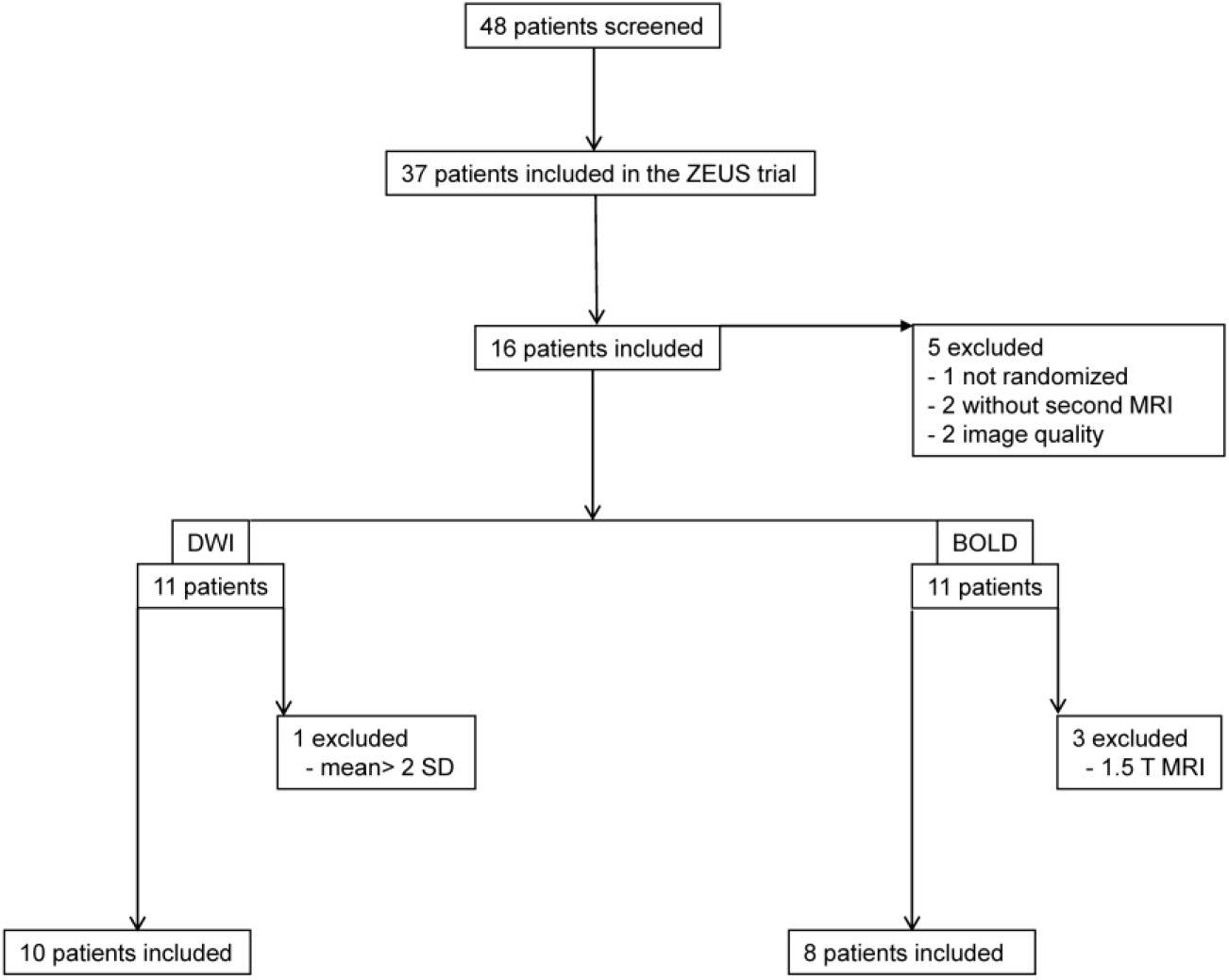
Flow chart. Patient screening and data exclusion. DWI (diffusion-weighted imaging); BOLD (blood oxygen level dependent-MRI).

#### 3.1.2. Patient characteristics

Baseline clinical characteristics are shown in Table 1. All of the patients were of Caucasian origin and mostly first transplant recipients with relatively preserved transplant function. Seven and nine subjects were randomized to the CsA and EVE groups respectively. There were more female subjects in the CsA group whereas living donor type was more frequent in the EVE group. Other variables were homogenously distributed among medication groups.

**Table 1.**
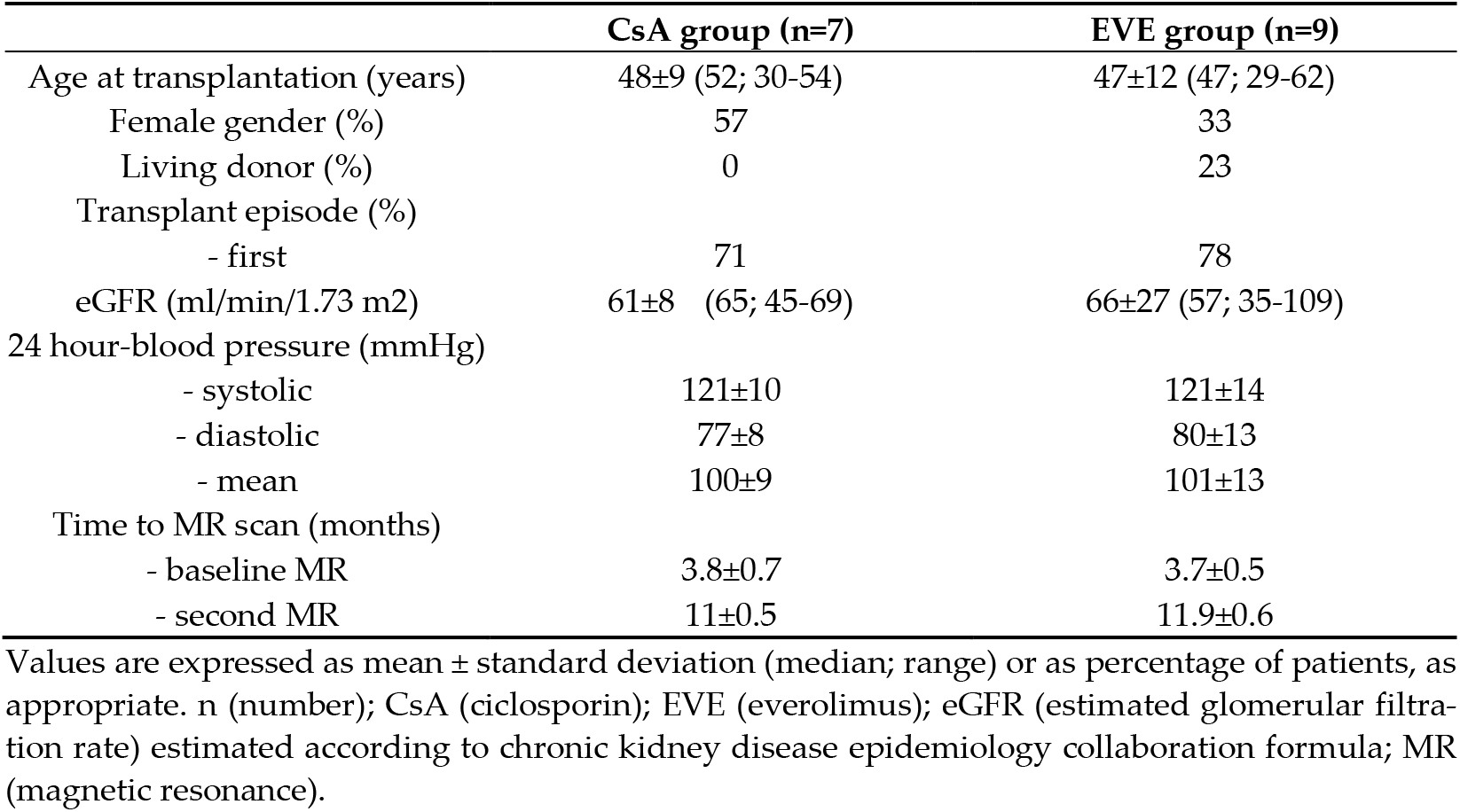
Baseline characteristics of the subjects (n=16).

### 3.2. Data quality

The MRI protocol including morphological sequences, DWI and BOLD could be performed in all of the patients at baseline and in 13 patients on study day 2. Subjects without second measurement were not included in the analysis (Figure 2). Data of two patients had to be excluded due to poor image quality resulting in analyzable data for 11 patients in each MR modality. In DWI, one patient had to be excluded because of outlier values (deviation >mean ± 2 SD); in BOLD, three patients were excluded due to field strength (1.5 T). A mean scanning time of one hour was met per session. DWI- and BOLD-derived mean values and SD ranges were roughly in line with previously reported values (Table 2) [17-20]. Low SD values confirmed measurement stability.

**Table 2.**
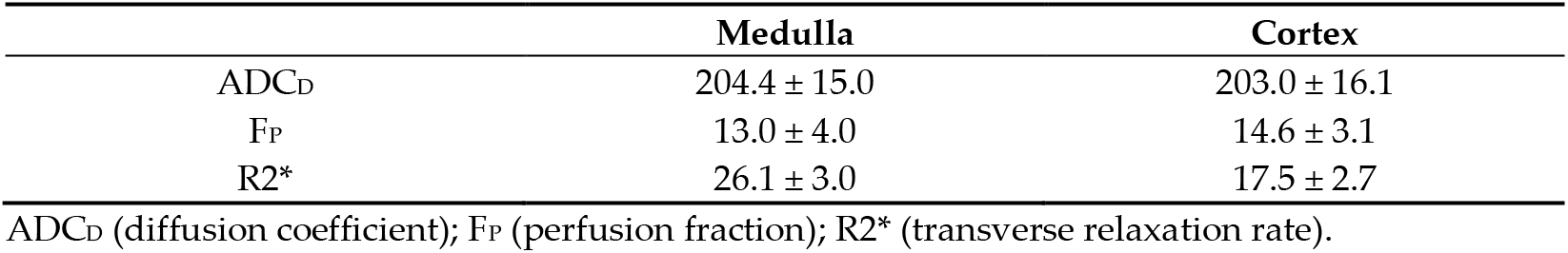
Overall mean values and standard deviations of diffusion-weighted imaging (DWI)- and blood oxygen level dependent (BOLD)-MRI-derived parameters.

### 3.3. Diffusion-weighted imaging

#### 3.3.1. Diffusion coefficient ADC_D_

Mean overall medullary and cortical values for the apparent diffusion coefficient ADC_D_ as a marker of pure diffusion are shown in Table 2 and Table 3. No cortico-medullary difference of the ADC_D_ was noted (*p*=0.65 at month 4.5, *p*=0.51 at month 12). In the CsA-treated group, medullary and cortical ADC_D_ values decreased in all but one subject from month 4.5 to month 12 (*p*=0.14 and *p*=0.14), whereas in the EVE-treated group, medullary and cortical ADC_D_ values increased from month 4.5 to month 12, reaching statistical significance for the medulla (*p*=0.046 and *p*= 0.12, respectively). At baseline, there was a tendency for higher ADC_D_ values in the group randomized to CsA as compared to the group randomized to EVE. In contrast, at month 12, ADC_D_ values were higher by trend in the EVE-treated group vs. the CsA-treated group (Table 3, Figure 3). This was the result of a significant difference of the mean change in ADC_D_ from month 4.5 to month 12 over time, which was negative in the CsA-treated group and positive in the EVE-treated group (Table 3, Figure 4). There were no differences in MCR ADC_D_ between time-points (*p*=0.47 for CsA-group; *p*=0.17 for EVE-group) or medication groups.

**Table 3.** DWI-derived pure diffusion coefficient according to medication group and time-point.

|  | CsA (n=4) | EVE (n=6) | *p*-value<sup>1</sup> |
| --- | --- | --- | --- |
| **Month 4.5** |  |  |  |
| ADC<sub>D</sub> medulla | 210.4±25.0 | 201.0±7.5 | 0.26 |
| ADC<sub>D</sub> cortex | 212.1±22.2 | 201.1±10.0 | 0.26 |
| MCR ADC<sub>D</sub> | 0.99±0.02 | 1.01±0.03 | 0.87 |
| **Month 12** |  |  |  |
| ADC<sub>D</sub> medulla | 195±14.5 | 210.0±12.2 | 0.17 |
| ADC<sub>D</sub> cortex | 193.5±20.0 | 206.3±13.7 | 0.26 |
| MCR ADC<sub>D</sub> | 1.01±0.05 | 1.02±0.04 | 0.78 |
| **Change month 4.5-12** |  |  |  |
| ΔADC<sub>D</sub> medulla | -15.4±18 | 9.0±7.6 | 0.019 |
| ΔADC<sub>D</sub> cortex | -18.6±23.8 | 6.2±7.3 | 0.038 |
| ΔMCR ADC<sub>D</sub> | 0.02±0.04 | 0.01±0.03 | 0.54 |

<sup>1</sup> Mann-Whitney U-test. Values are expressed as mean ± standard deviation. CsA (ciclosporin); EVE (everolimus); ADC<sub>D</sub> (diffusion coefficient) in 10<sup>-5</sup>mm<sup>2</sup>/s.

**Figure 3.**
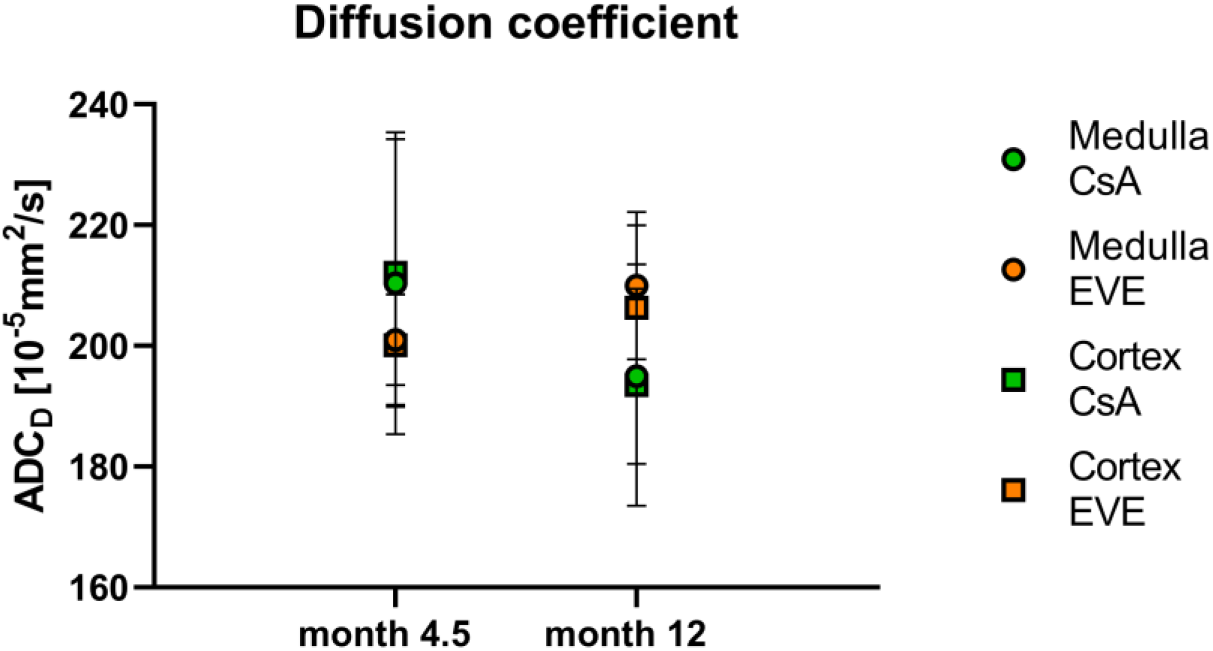
DWI-derived diffusion coefficient ADC_D_. Diffusion coefficient ADC_D_ according to medication group and time-point. CsA (ciclosporin); EVE (everolimus); ADC_D_ (diffusion coefficient).

**Figure 4.**
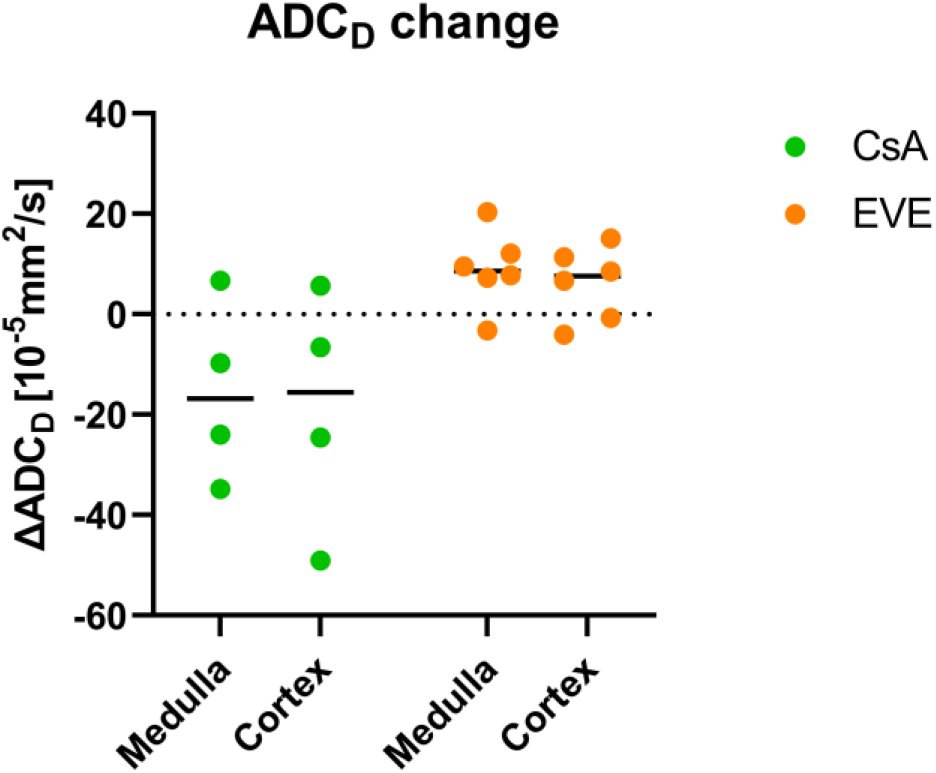
Change in DWI-derived diffusion coefficient. Change in diffusion coefficient ADC_D_ from month 4.5 to month 12. CsA (ciclosporin); EVE (everolimus); ADC_D_ (diffusion coefficient).

**Figure 5.**
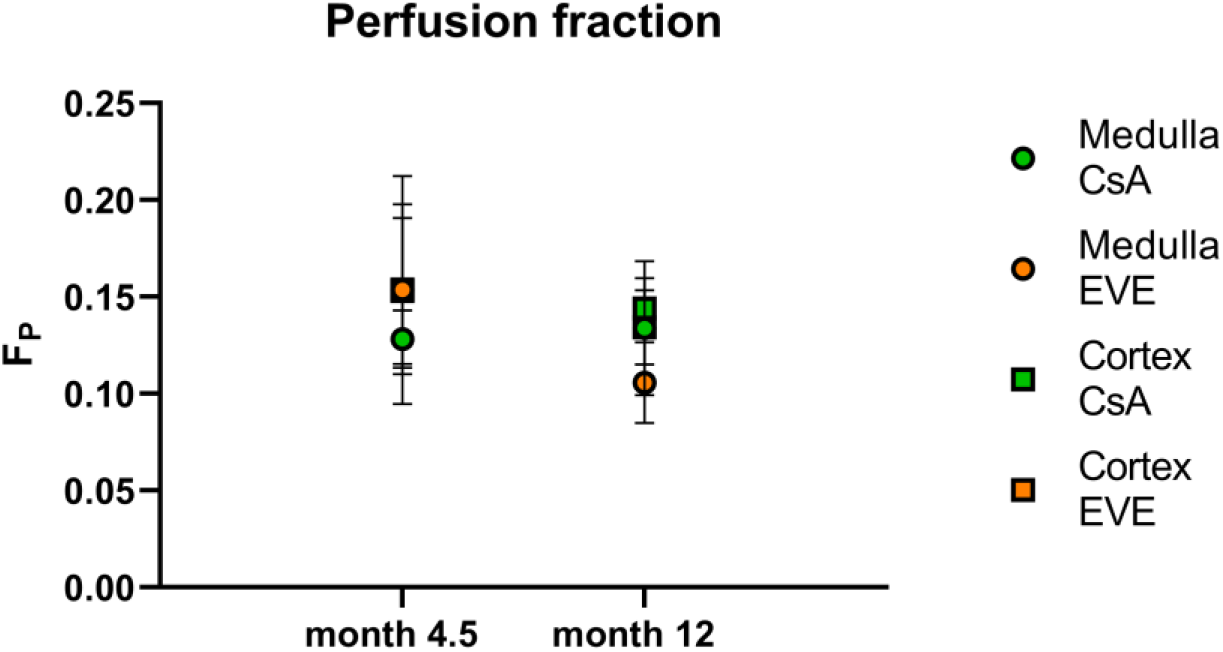
DWI-derived perfusion fraction. Perfusion fraction F_P_ according to medication group and time-point. CsA (ciclosporin); EVE (everolimus); F_P_ (perfusion fraction).

#### 3.3.2. Perfusion fraction F_P_

Mean medullary and cortical values for the fraction of perfusion F_P_ are shown in Table 2 and Table 4. As with ADC_D_, no significant cortico-medullary differences were found for F_P_ at both time-points (*p*=0.58 at month 4.5, *p*=0.059 at month 12). There was no difference in F_P_ or MCRF_P_ between medication groups at baseline nor at twelve months. Furthermore, no difference in the change over time of F_P_ or MCRF_P_ according to medication group was noted. Overall, no change was seen from baseline to month 12 across nor within medication groups in F_P_ in medulla (*p*= 0.17 for both groups; *p*=0.72 for the CsA-group; *p*=0.12 for the EVE-group), in cortex (*p*=0.29 for both groups; *p*=0.72 for the CsA-group; *p*=0.35 for the EVE-group) nor in MCRF_P_ (*p*=0.45 for both groups; *p*=0.47 for the CsA-group; *p*=0.25 for the EVE-group).

**Table 4.** DWI-derived perfusion fraction according to medication group and time-point.

|  | CsA (n=4) | EVE (n=6) | p-value<sup>1</sup> |
| --- | --- | --- | --- |
| **Month 4.5** |  |  |  |
| $F_P$ medulla | 12.8±1.5 | 15.4±5.9 | 0.26 |
| $F_P$ cortex | 15.3±3.8 | 15.4±4.4 | 1.00 |
| MCR $F_P$ | 0.9±0.3 | 1.0±0.2 | 0.87 |
| **Month 12** |  |  |  |
| $F_P$ medulla | 13.4±3.5 | 10.6±2.1 | 0.17 |
| $F_P$ cortex | 14.4±1.5 | 13.4±1.9 | 0.48 |
| MCR $F_P$ | 0.9±0.3 | 0.8±0.1 | 0.28 |
| **Change month 4.5-12** |  |  |  |
| $\Delta F_P$ medulla | 0.6±2.5 | -4.8±7.1 | 0.17 |
| $\Delta F_P$ cortex | -0.9±4.0 | -2.0±4.4 | 0.76 |
| $\Delta MCR F_P$ | 0.05±0.36 | -0.2±0.3 | 0.34 |

<sup>1</sup> Mann-Whitney U-test. Values are expressed as mean ± standard deviation. CsA (ciclosporin); EVE (everolimus); $F_P$ (perfusion fraction) in %.

### 3.4. Blood oxygen level dependent-imaging

Mean medullary and cortical values for the transverse relaxation rate R2* are shown in Table 2 and Table 5. As expected, significant cortico-medullary differences were shown at both time-points (*p*=0.012) with lower cortical R2* values, compatible with the known relative medullary hypoxia. Inverse trends were noted according to medication assignment including increasing medullary R2* values (i.e. reduced tissue oxygenation) in all of the CsA-treated vs. decreasing medullary and cortical R2* values in five and four of six EVE-treated patients respectively (Figure 6). However, the number and distribution of cases across groups at month 12 precluded formal statistical analysis. No changes in medullo-cortical distribution were seen between medication groups or time-points.

**Table 5.**
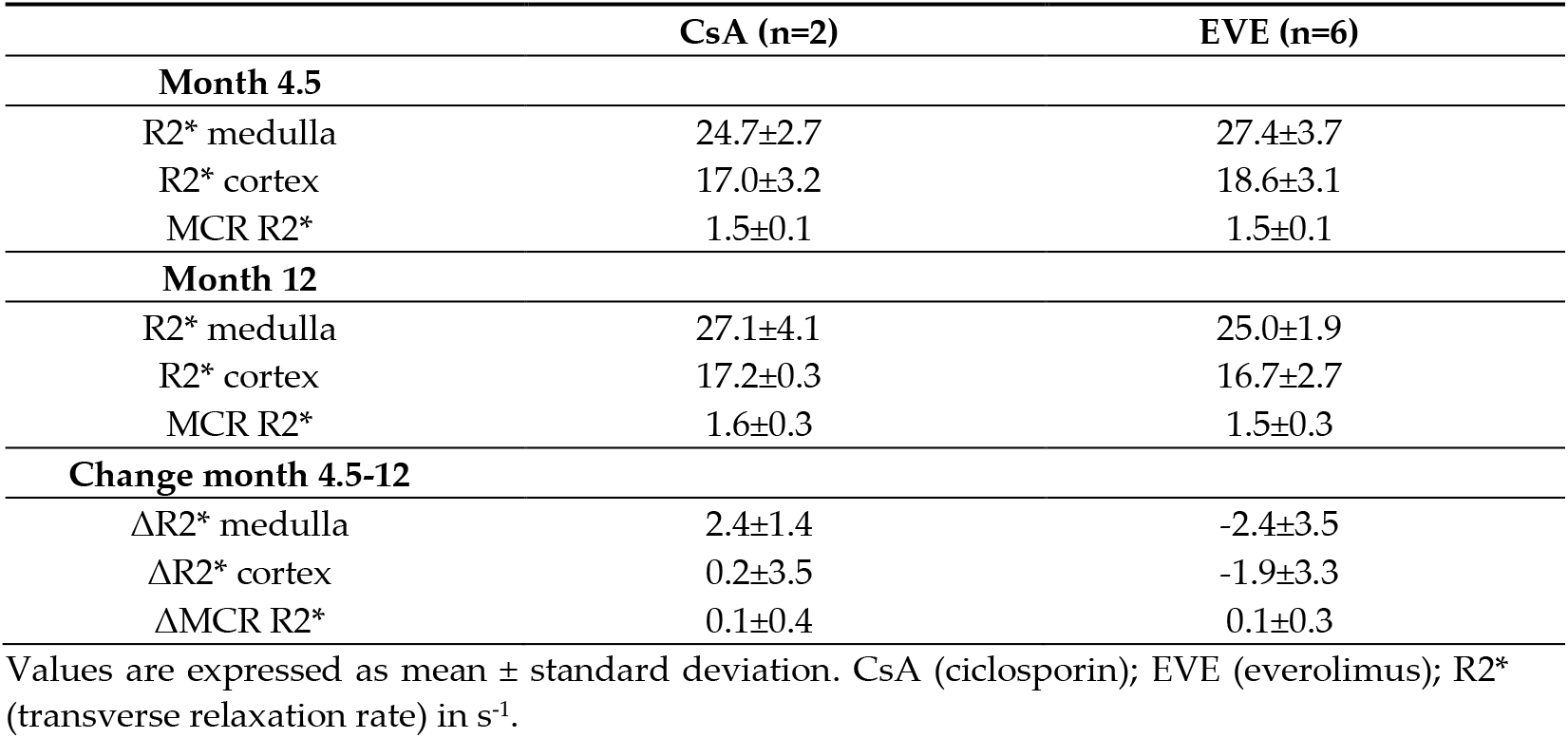
BOLD-derived transverse relaxation rate according to medication group and time-point.

**Figure 6.**
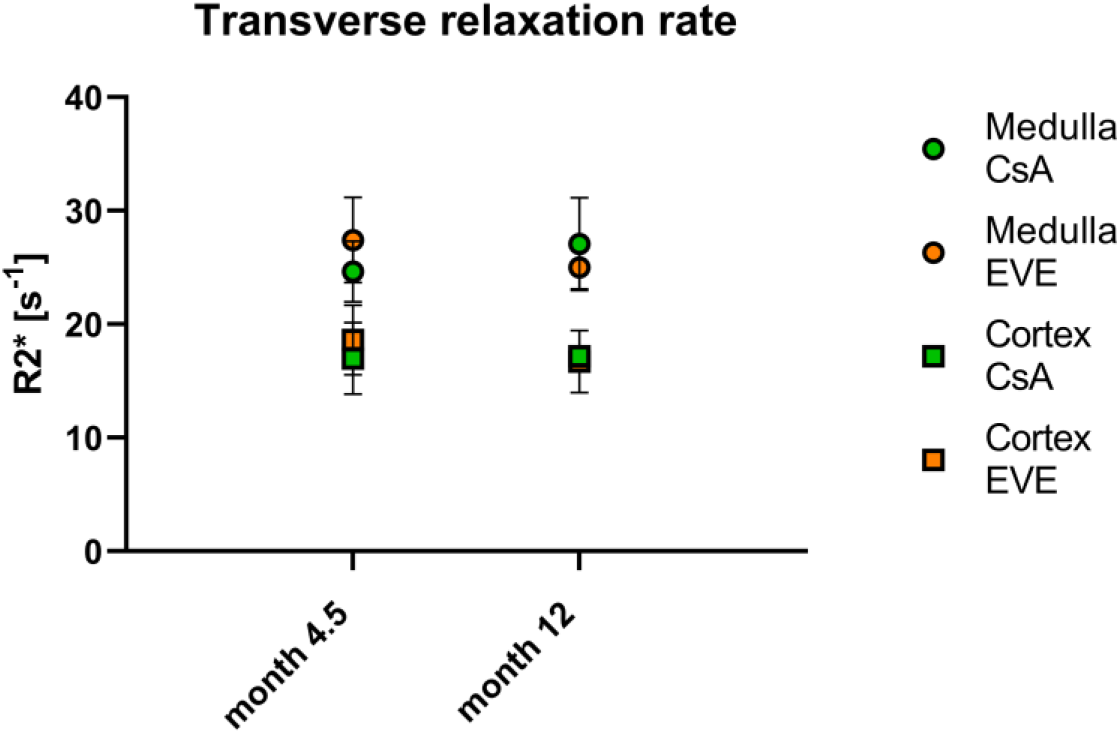
BOLD-derived transverse relaxation rate. Transverse relaxation rate R2* according to medication group and time-point. CsA (ciclosporin); EVE (everolimus); R2* (transverse relaxation rate).

### 3.5. Resistive indices

Mean resistive indices as measured by duplex ultrasound scan are shown in Table 6. Baseline RI values 4.5 months after transplantation corresponded to reference values in kidney grafts [21]. At baseline, measured RI were by trend higher in the group randomized to CsA than in the group randomized to EVE. At month 12 however, the difference reached statistical significance due to a tendency to greater increase of RI over time in the CsA-treated vs the EVE-treated group. No change was seen separately in the groups of CsA- and EVE-treated patients over time (*p*=0.35 and *p*=0.80 respectively).

**Table 6.** Mean resistive indices measured by duplex ultrasound according to medication group and time-point.

|  | CsA (n=6) | EVE (n=8) | p-value<sup>1</sup> |
| --- | --- | --- | --- |
| RI month 4.5 | 70.3±4.5 | 65.0±8.2 | 0.18 |
| RI month 12 | 71.7±4.5 | 65.8±7.3 | 0.04 |
| ΔRI (month 4.5-12) | 1.4±3.7 | 0.8±4.5 | 0.76 |

<sup>1</sup> Mann-Whitney U-test. Values are expressed as mean ± standard deviation. CsA (cyclosporin); EVE (everolimus); RI (resistive index).

### 3.6. Ambulatory blood pressure measurement

Mean ABPM-derived blood pressure values are shown in Table 7. There was a trend for increase in blood pressure from months 4.5 to 12 in the CsA-treated group whereas blood pressure values in the EVE-treated groups showed no evident change even though formal statistical analysis was not possible due to the number and distribution of cases at month 12. Conversely, nocturnal blood pressure dipping showed an increasing trend in the CsA-treated vs the EVE-treated group.

**Table 7.** Mean ABPM-derived blood pressure profile according to medication group and time-point.

|  | CsA (n=7)<sup>1</sup> | EVE (n=8) |
| --- | --- | --- |
| **Month 4.5** |  |  |
| BP sys | 121.3±10.3 | 120.9±13.5 |
| BP dia | 77.1±7.6 | 79.9±12.8 |
| BP mean | 99.6±8.6 | 100.9±13.2 |
| Dipping (sys/dia/mean) | 1±6.9/5±9.1/2.7±7.3 | 7.3±12.1/9.5±12.9/8.9±12.9 |

| Month 12 |  |  |
| --- | --- | --- |
| BP sys | 121.0±1.0 | 122.1±8.1 |
| BP dia | 77.3±2.1 | 79.8±4.8 |
| BP mean | 99.7±1.5 | 101.1±6.2 |
| Dipping (sys/dia/mean) | 4.3±10.2/6.3±11.4/5.0±10.6 | 3.5±11.9/9.3±11/6.0±11.1 |
| Change month 4.5-12 |  |  |
| ΔBP sys | 9.7±7.4 | 1.25±13.5 |
| ΔBP dia | 6.3±6.8 | -0.1±13.2 |
| ΔBP mean | 8±6.9 | 0.3±13.2 |
| ΔDipping (sys/dia/mean) | 3.7±16.7/0.3±22.6/2.0±18.5 | -3.8±10.0/-0.3±13.2/-2.9±10.9 |

<sup>1</sup>n=3 at month 12. Values are expressed as mean ± standard deviation. ABPM (ambulatory blood pressure monitoring); CsA (ciclosporin); EVE (everolimus); BP (blood pressure) in mmHg.

### 3.7. Laboratory parameters of graft function

Mean serum creatinine values and estimated glomerular filtration rates are shown in Table 8. At baseline, there was a tendency for lower eGFR in the group randomized to CsA. Whereas graft function was stable in the EVE-treated patients over time, it worsened by trend in the CsA-treated group. However, values at twelve months as well as changes from month 4.5 were not significantly different between medication groups.

**Table 8.** Mean serum creatinine values and estimated glomerular filtration rate according to medication group and time-point.

|  | CsA (n=7) | EVE (n=8) | p-value<sup>1</sup> |
| --- | --- | --- | --- |
| Month 4.5 |  |  |  |
| Serum creatinine | 112.0±21.0 | 12.01±40.0 | 0.46 |
| eGFR | 60.9±8.1 | 65.6±26.8 | 0.61 |
| Month 12 |  |  |  |
| Serum creatinine | 118.9±25.6 | 117±35.0 | 1.00 |
| eGFR | 57.1±11.0 | 65.5±19.7 | 0.40 |
| Change month 4.5-12 |  |  |  |
| ΔSerum creatinine | 6.9±17.2 | -4.4±22.3 | 0.46 |
| ΔeGFR | -3.7±10.3 | -0.1±16.5 | 0.87 |

<sup>1</sup> Mann-Whitney U-test. Values are expressed as mean ± standard deviation. CsA (ciclosporin); EVE (everolimus); eGFR (estimated glomerular filtration rate) according to chronic kidney disease epidemiology formula. Serum creatinine in μmol/L.

### 3.8. Protocol biopsies

The number of available biopseies was six and five at baseline as well as five and five at month 12 in the groups randomized to CsA and EVE respectively. No difference in the degree of IFTA (reaching from none to mild) or arteriolar hyalinosis (reaching from none to severe) was revealed between nor across medication groups (data not shown).

### 3.9. Correlation of fMRI parameters

To investigate medication-induced changes in fMRI parameters as well as correlations of fMRI parameters with selected clinical, biological and histological parameters, exploratory correlation testing was performed.

Changes in ADC_D_ over time were not correlated with changes in eGFR or histological parameters. Cortical ADC_D_ at twelve months correlated negatively with the resistive indices measured by DUS (τ=-0.511, *p*=0.040).

The perfusion fraction F_P_ correlated positively with eGFR at twelve months (τ=0.629, *p*= 0.012). A negative correlation was found between F_P_ and the diastolic and mean blood pressure by ABPM (τ=-0.584, *p*=0.020 and τ=-0.523, *p*=0.038 respectively).

No correlation of R2* with blood pressure, RI or graft function was noted.

## 4. Discussion

In this investigator-initiated study, we tested the hypothesis that diffusivity and perfusion as well as tissue oxygenation as measured by fMRI techniques differ between kidney transplant recipients included in the ZEUS trial randomized to continuing ciclosporin or to being switched to everolimus. The main findings are the following: (1) The mean change in medullary and cortical ADC_D_ significantly differed between both medication groups showing increase in EVE-treated patients while decreasing in CsA-treated patients. (2) Medullary ADC_D_ significantly increased after switch to everolimus. (3) Medullary and cortical R2* values showed inverse trends in medications groups with increasing values (i.e. reduced tissue oxygenation) in the CsA-treated group and decreasing values in the EVE-treated group.

First, ADC_D_ as a marker of perfusion-free diffusion decreased in patients maintained on CsA whereas it increased in patients converted to everolimus. The ADC_D_ has previously been shown to be reduced in various states of acute and chronic kidney diseases and to correlate to biological kidney function markers and interstitial fibrosis [9, 18, 22-25]. This course parallels the improvement of eGFR in EVE-treated patients reported in the ZEUS trial, which was by trend also observed in our sub-study population. Further, the decrease in diffusivity in CsA-treated patients might point to changes of tissue structure, possibly due to chronic nephrotoxicity patterns of calcineurin inhibitors with development of IFTA. Review of performed protocol biopsies did however not suggest increased proportion or severity of IFTA in the CsA-treated group even though the number of biopsies available was low and a sampling error cannot be excluded. In addition, the reversibility after 4.5 months of CsA treatment in the intervention group might suggest early injury or alternative, potentially functional mechanisms. Furthermore, an influence of the higher proportion of recipients of living donor kidneys in the EVE-treated group cannot be excluded.

Second, R2* values as marker of tissue oxygenation -if confounding factors are excluded-increased (lower tissue oxygenation) in CsA-maintained patients whereas it decreased (higher tissue oxygenation) in EVE-treated patients. This may suggest ameliorated graft tissue oxygenation under everolimus as compared to ciclosporin, possibly due to the expected vasoconstrictor effect of calcineurin inhibitors [14]. However, hematocrit represents a major confounding factor for BOLD-MRI with anemia leading to falsely-low R2* values and being expected as part of the everolimus side effect profile [26]. Despite lower hemoglobin values in the core as well as the current study in EVE-treated patients, hemoglobin values were not significantly different between both medication groups at both time-points [6].

The DWI-derived fraction of perfusion F_P_ showed no significant changes over time nor between medication groups. Studies in kidney grafts have suggested lower F_P_ values in cases of decreased graft function and acute rejection as well as to correlate to chronic tubulo-interstitial damage [18, 24, 27]. In our study, the number of cases might have prevented a significant finding. F_P_ correlated positively with eGFR at month 12, which corresponds to previous reports [18].

In this study, no cortico-medullary differences were found for ADC_D_ and F_P_. This is in accordance with previous studies from our center showing a loss of cortico-medullary differentiation over time in transplanted kidneys from living donors [18, 22, 23].

Other relevant findings include significantly higher resistive indices as measured by DUS in the CsA-treated group as compared to the EVE-treated group at month 12, possibly in the context of calcineurin vasculopathy. However, this difference, albeit non-significant, was present at baseline as well. In addition, higher resistive indices were associated with lower cortical ADC_D_ values at month 12. Similarly and expectedly, mean blood pressure values as measured by ABPM increased by trend in the CsA-treated group over time while remaining stable in the EVE-treated group.

There are important limitations to our study: First, the number of subjects available for final analysis was limited precluding formal statistical analysis of the BOLD data as well as multivariate analysis of clinical correlations. Second, for technical reasons, three patients underwent MRI at 1.5T as opposed to 3T for the other patients. These data were therefore excluded for BOLD-MRI. Lastly, post hoc correction for multiple comparisons was not carried out in this exploratory analysis of clinical correlations.

In conclusion, this prospective sub-study of the ZEUS trial suggests an impact of immunosuppressive regimen on fMRI parameters of the renal allograft.

## Data Availability

All data produced in the present study are available upon reasonable request to the authors

## Author Contributions

Conceptualization, U.E. and L.M.; methodology, P.V. and U.E.; software, P.V.; validation, P.V., J.C. and L.M.; formal analysis, J.C., P.V. and L.M.; investigation, U.E., P.V.; resources, U.E. and P.V.; data curation, U.E. and P.V.; writing—original draft preparation, L.M.; writing—review and editing, J.C., P.V., U.E., B.V.; visualization, J.C. and L.M.; supervision, B.V.; project administration, U.E.; funding acquisition, U.E. and P.V. All authors have read and agreed to the published version of the manuscript.

## Institutional Review Board Statement

The study was conducted in accordance with the Declaration of Helsinki, and approved by the Ethics Committee of the Canton of Bern, Switzerland (protocol code 2004/213).

## Informed Consent Statement

Informed consent was obtained from all subjects involved in the study.

## Data Availability Statement

The data presented in this study are available on request from the corresponding author. The data are not publicly available due to privacy reasons.

## Acknowledgments

We thank all patients who participated in this study. The results of this study have been presented at the 57^th^ Congress of the European Renal Association-European Dialysis and Transplant Association.

